# Effect of hot zone infection outbreaks on the dynamics of SARS-CoV-2 spread in the community at large

**DOI:** 10.1101/2020.11.23.20237172

**Authors:** Dominik Wodarz, Natalia L. Komarova, Luis M. Schang

## Abstract

Transmission of SARS-CoV-2 appears especially effective in “hot zone” locations where individuals interact in close proximity. We present mathematical models describing two types of hot zones. First, we consider a metapopulation model of infection spread where transmission hot zones are explicitly described by independent demes in which the same people repeatedly interact (referred to as “static” hot zones, e.g. nursing homes, food processing plants, prisons, etc.). These are assumed to exists in addition to a “community at large” compartment in which virus transmission is less effective. This model yields a number of predictions that are relevant to interpreting epidemiological patterns in COVID19 data. Even if the rate of community virus spread is assumed to be relatively slow, outbreaks in hot zones can temporarily accelerate initial community virus growth, which can lead to an overestimation of the viral reproduction number in the general population. Further, the model suggests that hot zones are a reservoir enabling the prolonged persistence of the virus at “infection plateaus” following implementation of non-pharmaceutical interventions, which has been frequently observed in data. The second model considers “dynamic” hot zones, which can repeatedly form by drawing random individuals from the community, and subsequently dissolve (e.g. restaurants, bars, movie theaters). While dynamic hot zones can accelerate the average rate of community virus spread and can provide opportunities for targeted interventions, they do not predict the occurrence of infection plateaus or other atypical epidemiological dynamics. The models therefore identify two types of transmission hot zones with very different effects on the infection dynamics, which warrants further epidemiological investigations.

## 1. Introduction

Infection spread through human communities has been studied extensively with mathematical models, which has resulted in a wealth of knowledge about the principles that drive the epidemiological dynamics of infectious diseases [1, 2]. At the core of this work are models that take into account populations of susceptible and infected individuals (SIS models), or more complex models that further take into account recovered/immune individuals (SIR models). Many models assume well-mixed populations, although the inclusion of age structure [1, 2], spatial structure [3-5], and contact networks [6-8] have been equally important.

A common assumption is that infections spread through the community at large by people coming into contact with each other socially and at work during everyday activities, which is a good assumption for many respiratory infections such as influenza. Variations of this scheme have been explored, such as the impact of super-spreaders [9], which are individuals who contribute a disproportionately large amount to infection spread in the community. Another deviation from the standard infection model assumptions likely applies to the SARS-CoV-2 pandemic: It appears that this infection spreads most efficiently in certain settings where a larger number of people interact for a prolonged period of time indoors (such as in hospitals, assisted living facilities, food processing plants, worker dormitories, prisons, bars, and restaurants) [10, 11]. In contrast, other, every day encounters between people in the community at large are less efficient at spreading the virus, especially if those occur outdoors. We call the places in which the infection spreads more efficiently “transmission hot zones”. In hot zones, transmission might be facilitated by the repeated interaction among a small subset of the population, which can amplify the virus within this group of people [12]. For example, in health care settings, patients with existing medical conditions frequently visit, in some cases daily, and medical staff interact daily. Similarly, in food processing plants or prisons, the same people converge daily, opening up the possibility for efficient viral amplification through repeated interactions in closed settings.

In previous work [13], we analyzed a mathematical model based on ordinary differential equations in which the virus was transmitted through two pathways: through the community at large, in which virus dose was assumed to be low and disease was likely mild, and through a hot zone pathway, in which virus dose was assumed to be relatively high, resulting in a greater chance of severe infections. Analysis concentrated on how these assumptions influenced the basic reproductive number of the virus. This work suggested the possibility that infection spread can hinge upon high-dose hot zone transmission, even though most infections were predicted to be mild and occur in the community at large [13]. Here we develop the concept of hot zone transmission further by explicitly modeling hot zone formation and virus transmission within hot zones. We describe models of two types of hot zones, which we call “static” and “dynamic” hot zones.

“Static hot zones” are given in the form of a patch or deme or metapopulation model. In this model, the community at large is described by one compartment, which has the largest population size. Individual hot zones are described by an array of patches, each of which is characterized by a relatively small population size, corresponding to subpopulations that regularly and repeatedly meet in a given hot zone, such as a nursing home, food processing plant, or prison. We find that hot zone transmission can serve as an infection reservoir that can prevent the continued decline of infection levels either following natural infection spread, or following the implementation of non-pharmaceutical intervention measures. Due to these hot zone dynamics the model can reproduce atypical epidemiological patterns found in COVID19 case data, such as prolonged infection plateaus during interventions [14] or sub-exponential growth patterns [15]. The dynamics predicted by the model further point towards complexities in estimating the basic reproduction number from epidemiological data that document infection spread in the overall community.

Next we consider “dynamic hot zones”, which are assumed to form spontaneously by randomly drawing individuals from the general population, and which are assumed to subsequently dissolve. This corresponds to settings where people randomly gather for a relatively short period of time and then disperse again, such as restaurants, bars, or movie theaters. We find that this model has very different properties compared to the static hot zone model, and that it cannot account for the same unique dynamical patterns. The model predicts standard epidemiological growth patterns, although dynamic hot zones accelerate the average rate of virus spread and thus present opportunities for targeted interventions. This points towards the presence of two types of hot zones with vastly different effects on the dynamics of infection spread.

## 2. Static hot zones, expressed by deme/patch models

We describe a mathematical model that takes into account different compartments/demes/patches, where the dynamics in each compartment are given by ordinary differential equations. The model includes one “community at large” compartment, and N hot zone compartments. The infection spreads separately in the community at large and in each hot zone according to modified SIR equations. The two types of locations, however, interact in the sense that members of the community at large can bring virus to the hot zones and seed infections there, and infected individuals in the hot zones can pass the virus on to members of the community at large. These principles are illustrated schematically in Figure 1. The assumption implicit in the compartment/deme description of the hot zones is that the hot zones are “static” in the sense that the same group of people spends extended periods of time there and interacts repeatedly in closed quarters, somewhat separate from the community at large. Examples are prisons, nursing homes, hospitals, etc. The equations underlying the model are summarized as follows.

**Figure 1.**
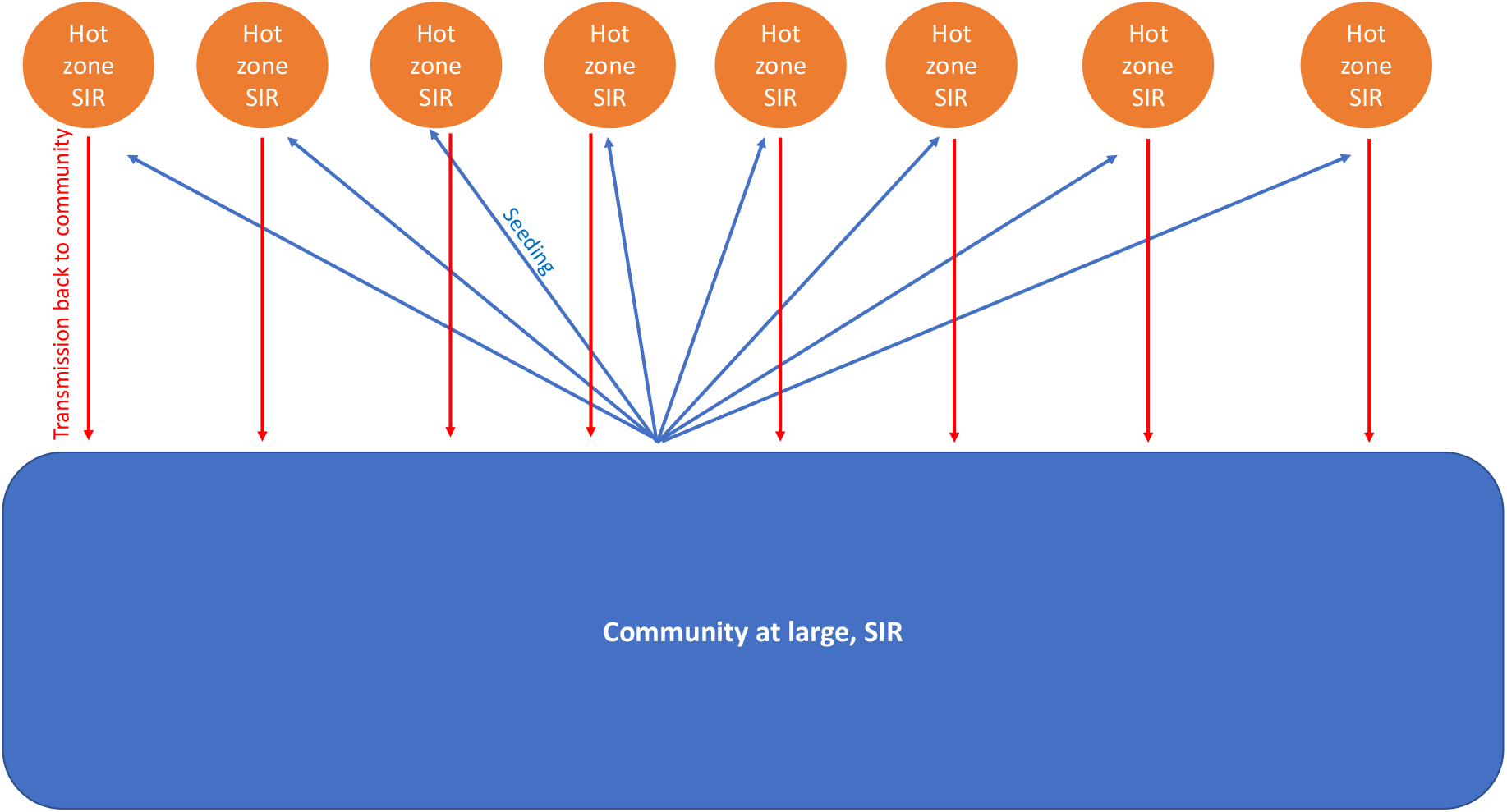
Schematic representation of model assumptions.

The time-evolution of infection spread in the community at large is described by the following ordinary differential equations:

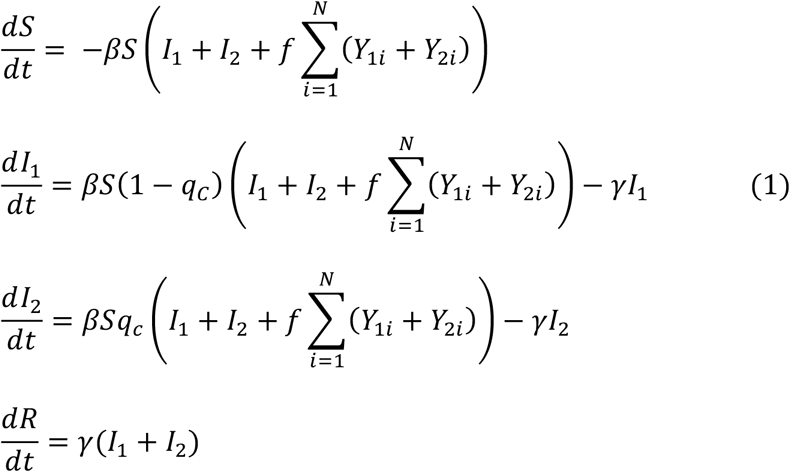

The population of susceptible individuals is denoted by S, the population of mildly and severely infected individuals is denoted by I_1_ and I_2_, respectively, and the population of removed individuals (recovery and dead) is given by R. The populations Y_1i_ and Y_2i_ represent mildly and severely infected people in the hot zones *i*=1..N. Susceptible individuals become infected by members of the community with a rate *β*, and by people from the hot zones with a rate *fβ*. Upon infection, severe disease occurs with a probability q_c_, while mild disease occurs with a probability 1-*q*_*c*_, where *q*_*c*_ = *(I*_*1*_+*I*_*2*_*)/K*_*c*_ if *I*_*1*_+*I*_*2*_ < *K*_*c*_, and *q=1* otherwise (*K*_*c*_ is equal or less to the total community population size).

In other words, the proportion of severe infections increases with virus prevalence, and all infections become severe if the fraction of infected individuals in the population reaches a relatively high threshold level. This captures observations from coronavirus transmissions, which indicate higher disease severity in people exposed to larger viral doses [16-19]. Infected individuals are assumed to recover or die with a rate *γ*.

The infection dynamics in the hot zones are described by similar ordinary differential equations, which are given as follows:

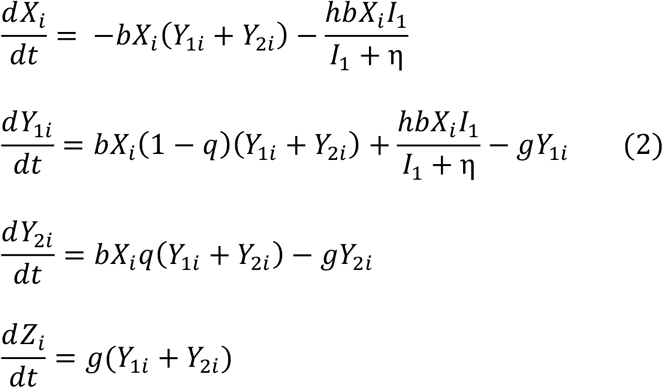

The subscript *i* denotes the identity of the hot zone, where *i=1*..*N*. In these compartments, susceptible individuals are given by X, mildly and severely infected individuals by Y_1_ and Y_2_, respectively, and removed individuals by Z. The population size in each hot zone is assumed to be very small compared to that in the community at large. The processes that are modeled in the hot zones are similar to the ones in the community at large. Thus, infection events occur with a rate b and recovery/death occurs with a rate g. The probability of severe infection in the hot zone, q_h_, is defined in the same way as in the community at large, *q*_*h*_ = *(Y*_*1*_+*Y*_*2*_*)/K*_*h*_ if *Y*_*1*_+*Y*_*2*_ < *K*_*h*_, and *q=1* otherwise (where *K*_*h*_ is equal or less to the total population size in the particular hot zone). In addition to these zone-specific infection processes, susceptible individuals in the hot zone can become infected by contact with members of the community at large with a rate hbX_i_I_1_/(I_1_+ξ), which can seed a hot zone. We assume that this can only occur through contact with mildly infected individuals from the community at large, since severely infected individuals are unlikely to enter the hot zones. Moreover, we assume that this infection rate is a saturating function of the number of mildly infected individuals in the community at large. The community compartment contains many more individuals than a hot zone. It would thus be unrealistic to assume that the number of infected people entering the hot zone is directly proportional to the number of infected individuals in the community. An analysis of a simplified ODE model is presented in the Supplementary Materials, including R_0_ calculations and solution structure.

An unrealistic consequence of formulating the model as a set of ordinary differential equations is that all hot zones become simultaneously seeded once there is any amount of virus in the community at large. This changes if the model is re-formulated stochastically through Gillespie simulations [20] of the ODEs. Now, entry of the infection into the hot zones is probabilistic, and an increasing number of hot zones can be successively seeded as the infection spreads in the community at large. This can lead to very different dynamics, as is demonstrated in the Supplementary Materials. It turns out, however, that this difference in behavior between the deterministic ODEs and the more realistic stochastic simulations is only due to the stochastic seeding of the hot zones. For the sake of simplicity, we therefore simulated the dynamics largely in a deterministic way by using the above-described ODEs, but without the saturated infection term that describes virus transmission from the community at large to susceptibles in the hot zones. Instead, we assumed that at each discrete unit of time, every hot zone has a probability pI_1_/(I_1_+ξ) to pick up the virus from the community at large, which is simulated by setting the percent infected in that hot zone to a low level if it was previously devoid of virus. This algorithm preserves the properties that arise from the stochasticity of the dynamics without having to simulate a full stochastic system, which is characterized by a higher variability and by a relatively high computational cost.

### 2.1. Basic dynamics: community at large vs hot zones

We investigate the effect of hot zones on the infection spread dynamics. We start by assuming that the basic reproductive number of the virus within the community at large is the same as in each hot zone, with R_0_=2.5 [21, 22]. We start computer simulations with a small percent of people infected in the community at large, and infection-free hot zones.

Figure 2A shows the dynamics in the absence of any interventions. In the community at large, we observe typical infection spread dynamics: The infection grows to a peak, and then declines to extinction. Hot zones are seeded during this process, and the infection in the hot zones eventually also goes extinct. We observe oscillatory dynamics in the hot zone infection levels, because the infection spreads through individual hot zones and subsequently declines towards extinction before the next hot zones are seeded. Since the infection goes extinct in the community at large, seeding of additional hot zones comes to an end as well. Figure 2B-D show the effect of non-pharmaceutical interventions, which are implemented by reducing the community infection rate, β, 3-fold. Figure 2B shows very early intervention. The reduction in infectivity occurs when the total percent of infected individuals in the community reaches 0.01%. We now observe a prolonged phase during which infection levels remain relatively steady, which can be called a “plateau phase”. Hot zones are seeded from the community, and the cross-infection between hot zones and the community maintains the infection over a longer period of time. In this simulation, the hot zones act as infection reservoirs that fuel the longer-term persistence. Without the hot zones, the infection goes extinct following the onset of the intervention (not shown). Infection levels fluctuate around a relatively steady value because the presence of the virus in individual hot zones is temporary, eventually resulting in virus extinction in that hot zone. Over time, however, the infection can be seeded in additional hot zones and the balance between local infection extinction and seeding of new hot zones drives the fluctuation around steady levels, until all hot zones have already been infected. Figure 2C shows a similar simulation, but the intervention is started when community infection levels reach 1%. Now, a more pronounced infection decline is observed after the intervention is put in place, but a steady plateau phase is again reached, similar to the previous case. Figure 2D shows a simulation where the intervention was implemented when total community infection levels reached 15%. Now, the dynamics are similar compared to those without intervention (panel A). The infection goes extinct relatively fast, and the hot zones cannot maintain a longer-term plateau phase. The reason is that the more extensive infection spread prior to the intervention leads to a more pronounced reduction in the percentage of susceptible individuals, and now the community infections from hot zones are not sufficient to maintain community spread. Instead the infection declines in the community, which results in reduced seeding of hot zones, which prevents longer term persistence.

**Figure 2.**
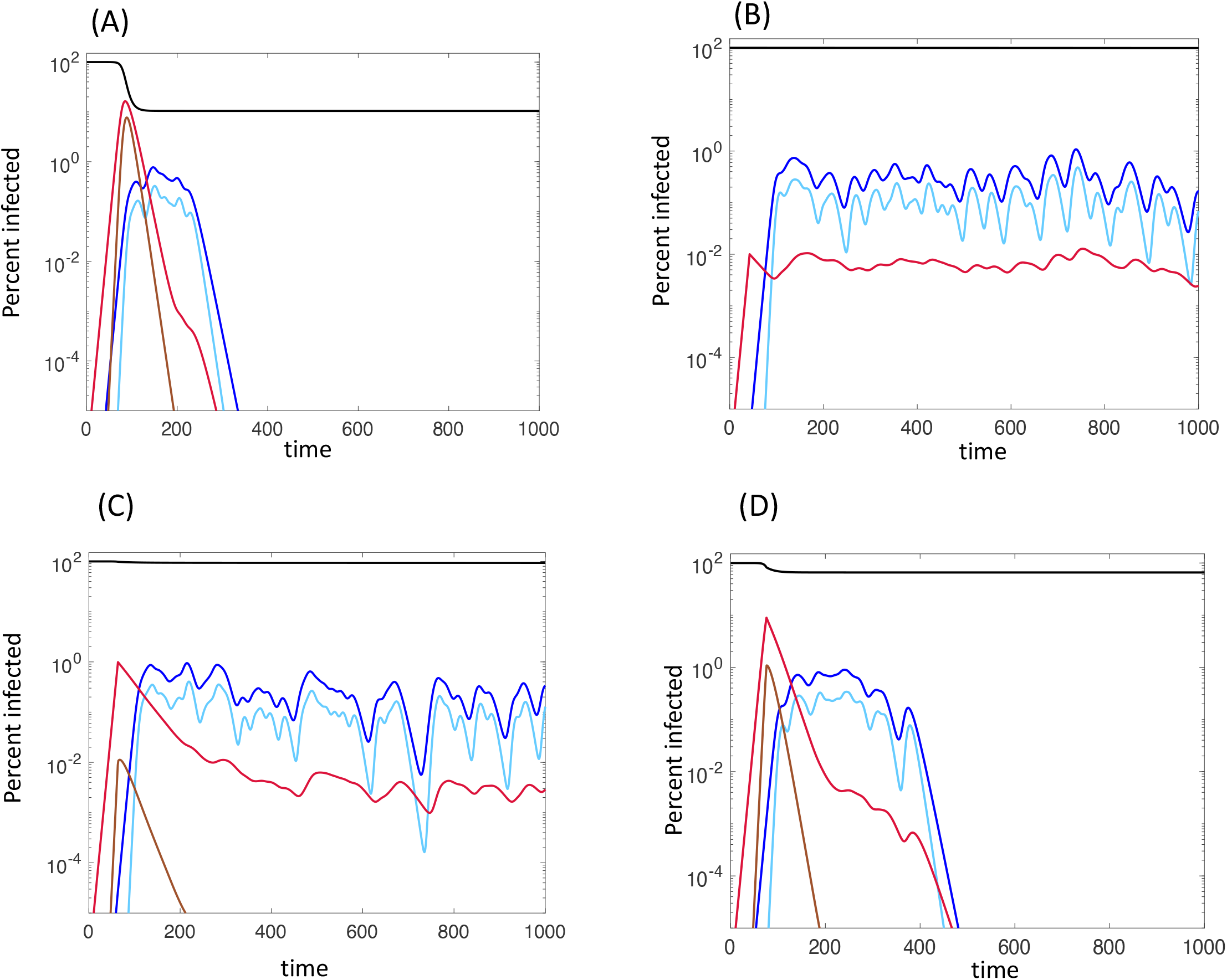
Effect of hot zones on the dynamics of infection spread. R0=2.5 for both the community at large and the hot zones is assumed, and the infection is simulated starting from a small amount of infecteds in the community at large. The percent of infected people are plotted. Black shows the percent of uninfected individuals in the community at large. Red and brown show mildly and severely infected individuals in the community at large. Dark and light blue show the percent of all mildly and severely infected individuals across all the hot zones. (A) Infection dynamics without interventions. (B) Intervention: the infectivity in the community at large is reduced 3-fold when the total percent of infected individuals in this compartment reaches 0.01%. (C) Same intervention, implemented when the total percent of infected individuals in the community reaches 1%. (D) Same intervention, implemented when the total percent of infected individuals in the community reaches 15%. Parameters were chosen as follows. S_0_=1000, β=0.36×10^−3^, γ=1/7, f=0.03, X_0_=1, b=0.36, g=1/7, p=0.001, f=0.03, ξ=0.01, K_c_=600, K_h_=0.6 N_zones_=100.

### 2.2. Slow rate of virus spread in the community at large compared to hot zones

Now we assume that the virus has a relatively low potential to spread through the community at large, with a basic reproductive number R_0_=1.25. In contrast, a relatively large basic reproductive number R_0_=3.75 is assumed for the hot zones, since these environments likely facilitate viral spread. Figure 3A shows a simulation of the dynamics in the absence of interventions. The hot zone dynamics are similar as described above, but the infection spread in the community at large shows interesting and atypical patterns in the current scenario. During the initial phase, a relatively slow spread of mild infections is observed (red line), due to the assumed low reproduction potential of the virus in the community at large. This might correspond to a slow, silent amplification of the infection. As the hot zones are seeded, the rate at which mild infections grow in the community at large increases rapidly and approximates the rate at which the virus spreads in the hot zones, which is significantly faster. We note that although the virus reproduction potential in the community is relatively low, community spread is driven by the hot zones during this phase, contributing to the accelerated infection growth in the community. At this time, severe infections in the community start to reach higher levels as well (brown line). This might correspond to the time when the infection is detected by tests in the community at large. As the infection levels stabilize in the hot zones (due to the balance between local virus extinction and seeding of new hot zones), the rate of virus spread in the community at large slows down again as the community infection levels continue to grow towards a peak and subsequently decline. Because we assumed a relatively low basic reproductive number of the virus in the community at large, the rate of infection decline post peak is also slower. This allows for continued seeding of further hot zones, which in turn boosts further infection spread in the community. This results in a quasi-steady state, during which infection levels fluctuate around a constant level.

**Figure 3.**
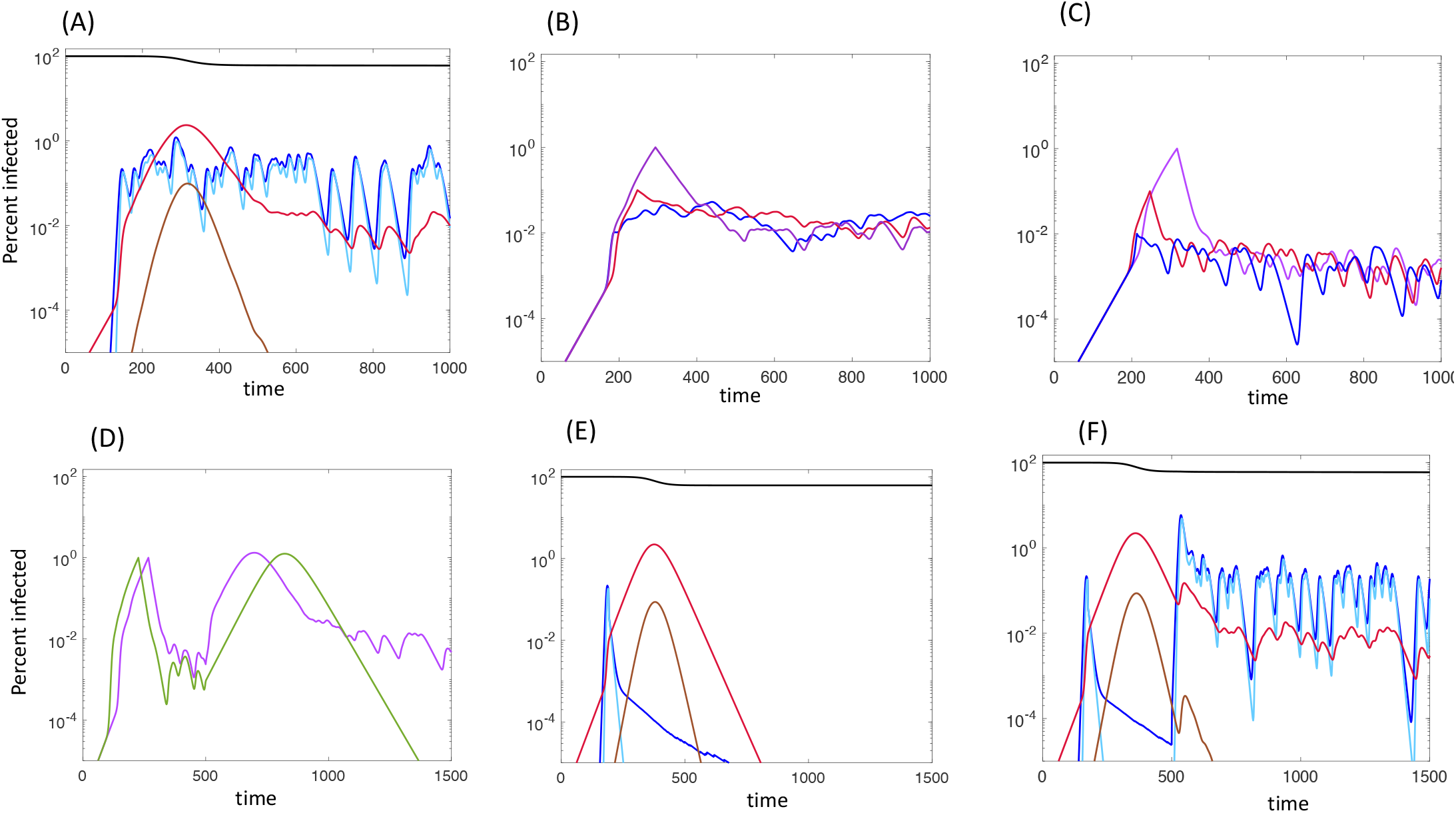
Model simulation assuming that in the community at large, R_0_=1.25, and in the hot zones, R_0_=3.75. (A) Dynamics without interventions. The percent of infected people are plotted. Black shows the percent of uninfected individuals in the community at large. Red and brown show mildly and severely infected individuals in the community at large. Dark and light blue show the percent of all mildly and severely infected individuals across all the hot zones. (B) Community interventions starting at different times, characterized by a 1.5-fold reduction of the community infectivity, β. The total percent of infected individuals is plotted. (C) Community interventions starting at different times, characterized by a 3-fold reduction of the community infectivity, β. The total percent of infected individuals is plotted. (D) Implementation of community interventions followed by their cessation. During intervention in the community at large, the rate of community infection β, is reduced 3-fold. We assumed a relatively late start of interventions here, when total community infection levels reached 1% of the total population. The green line shows the dynamics in the community at large without the existence of hot zones, while the purple line shows a simulation under the standard assumption that hot zones exist. (E) Implementation of hot zone interventions, in the absence of interventions in the community at large. Color codes are the same as in panel A. During hot zone interventions, the hot zone population size is reduced by 75%, and this was initiated relatively early when total community infections reach 0.01% of the total population. (F) Implementation of hot zone interventions (without interventions in the community at large), with subsequent cessation. Color codes are the same as in panel A. Parameters were chosen as follows. S_0_=1000, β=0.18×10^−3^, γ=1/7, f=0.1, X_0_=1, b=0.54, g=1/7, p=0.001, ξ=0.01, K_c_=600, K_h_=0.6 N_zones_=100.

We would like to point out two characteristics of these dynamics. First, infection spread does not follow a straightforward exponential law, but is more complex. The mild infections transition from slow to faster, back to slower growth. Consequently, the severe infections also grow sub-exponentially. Second, a fast infection spread can be observed in the community at large during the time frame when community spread is promoted by expanding infection levels in the hot zones. This can contribute to an estimation of a community R_0_ value that is between 2-3, although in reality it is assumed to be 1.25 in this simulation. Because the community R_0_ is assumed to be relatively low, the infection also peaks at low levels (in the absence of interventions), when only about 20% of the total population has been infected. This would be considered unexpected if the community R_0_ was estimated to lie between 2 and 3.

Figure 3B and C show simulations that include non-pharmaceutical interventions, with the different lines varying in the time when the interventions were initiated. Panels B and C differ in the intensity of the interventions. The community infection rate β was reduced 1.5-fold in panel A, and 3-fold in panel B. We observe that regardless of the time at which interventions are initiated, the dynamics converge to fluctuating around the same quasi steady state level during the interventions, and that this level is lower for stronger interventions (compare panels B and C). Eventually, the infection will go extinct once no more hot zones are susceptible to outbreaks (not shown).

Figure 3D shows simulations of non-pharmaceutical interventions in the community at large, with subsequent relaxation of these interventions. The purple line corresponds to our standard assumptions, and the green line shows a corresponding simulation where upon relaxation of community interventions, hot zone transmission is eliminated. While the predicted second waves in the community at large have a similar magnitude in the two cases, the infection declines towards faster extinction in the absence of hot zone transmission (green line), while it stabilizes and continues to be maintained for a certain period of time if hot zone transmission remains in place (purple line). This again points towards the hot zones as a reservoir compartment that can maintain infections over prolonged durations.

Figure 3E shows a simulation in which no interventions are put in place in the community at large, but interventions are implemented in the hot zones. The hot zone intervention was modeled by reducing the population size in each hot zone by 75%, which results in an effective hot zone reproduction number that is lower than one, and prevents successful infection spread in each hot zone. Following the initiation of this intervention, hot zone infection levels decline, but infection spread in the community at large continues for a while, at a rate determined by the assumed low infectivity in the community. The infection peaks due to community herd immunity when about 20% of this population has been infected (due to the assumed low basic reproductive number in the community), and subsequently declines towards extinction. Figure 3F shows a similar simulation, but where hot zone interventions are reversed before the infection has gone extinct. This results in renewed successful hot zone seeding, with only a small blip in community transmission, driven by the additional virus transmission from hot zones. The infection levels in the community at large, however, subsequently continue to fluctuate around steady levels (rather than decline further), due to the infection-maintaining role of the hot zones.

### 2.3. The rate of community spread as a function of viral dose

The last section showed that a slower rate of virus spread in the community at large compared to that in the hot zones can result in interesting dynamics that are relevant to SARS-CoV-2 epidemiology. One consequence of this model assumption was that in the absence of interventions, community infection levels peaked when only 20% of the whole community population had become infected. This low herd immunity threshold is a direct result of the assumed low viral reproduction number in the community at large. Data, however, indicate that the virus might spread more extensively, e.g. up to 40%-60% cumulative infection levels in the Amazon in Brazil [23]. Here, we consider a modification of the model explored in the previous section: We will assume that severely infected individuals transmit the virus with a larger rate than mildly infected ones. As in the previous section, the probability of severe infection increases with the total amount of virus to which people are exposed (i.e. with the total number of infecteds in the population, both in the community at large and in the hot zones). Consequently, as the infection levels grow, the average transmission rate increases due to the presence of a higher number of severely infected individuals. Therefore, at low infection prevalence, the virus spread rate in the community at large is relatively slow as in the previous section, but average viral spread accelerates as the infection levels grow.

During the initial phases of spread, we observe similar dynamics as in the previous version of the model (Figure 4Ai), because at low infection prevalence, severe disease is rare and the infection spreads slowly. The spread begins in the community at large, and is initially slow and only consists of mild cases. Over time, hot zones are seeded, which speeds up the growth of infection levels in the community at large, and results in the emergence of severe disease. The reason for these dynamics is the same as in the previous section. Once the infections in the hot zones start to stabilize (due to a balance between virus extinction in some hot zones and seeding of additional ones), however, the rate of community virus spread slows down to a lesser degree compared to the previous model. The reason is that hot zone spread resulted in a strong amplification of virus in the community and the occurrence of more severe disease, and this is assumed to increase the transmission rate in the current version of the model. It follows that a higher peak infection level is observed in the community at large compared to the previous model, and post peak, cumulative infection levels range between 40%-60%, which is markedly higher than in the model analyzed in the last section. Therefore, in the current model version, all key observations about the dynamics pointed out in the last section remain robust, with the exception that now herd immunity sets in at significantly higher cumulative infection levels, which might be more realistic. Figure 4Aii shows the total percent infections summed over the community at large and all hot zones. Similar to the dynamics in the individual sub-populations, we observe that once the dynamics stabilize post-peak due to hot zone transmission, temporal oscillations are observed in total infection levels, which are again due to the balance between virus extinction in individual hot zones, and seeding of new hot zones. Similar patterns are found when only severe infections are summed up over the community at large and all hot zones (Figure 4Aiii).

**Figure 4.**
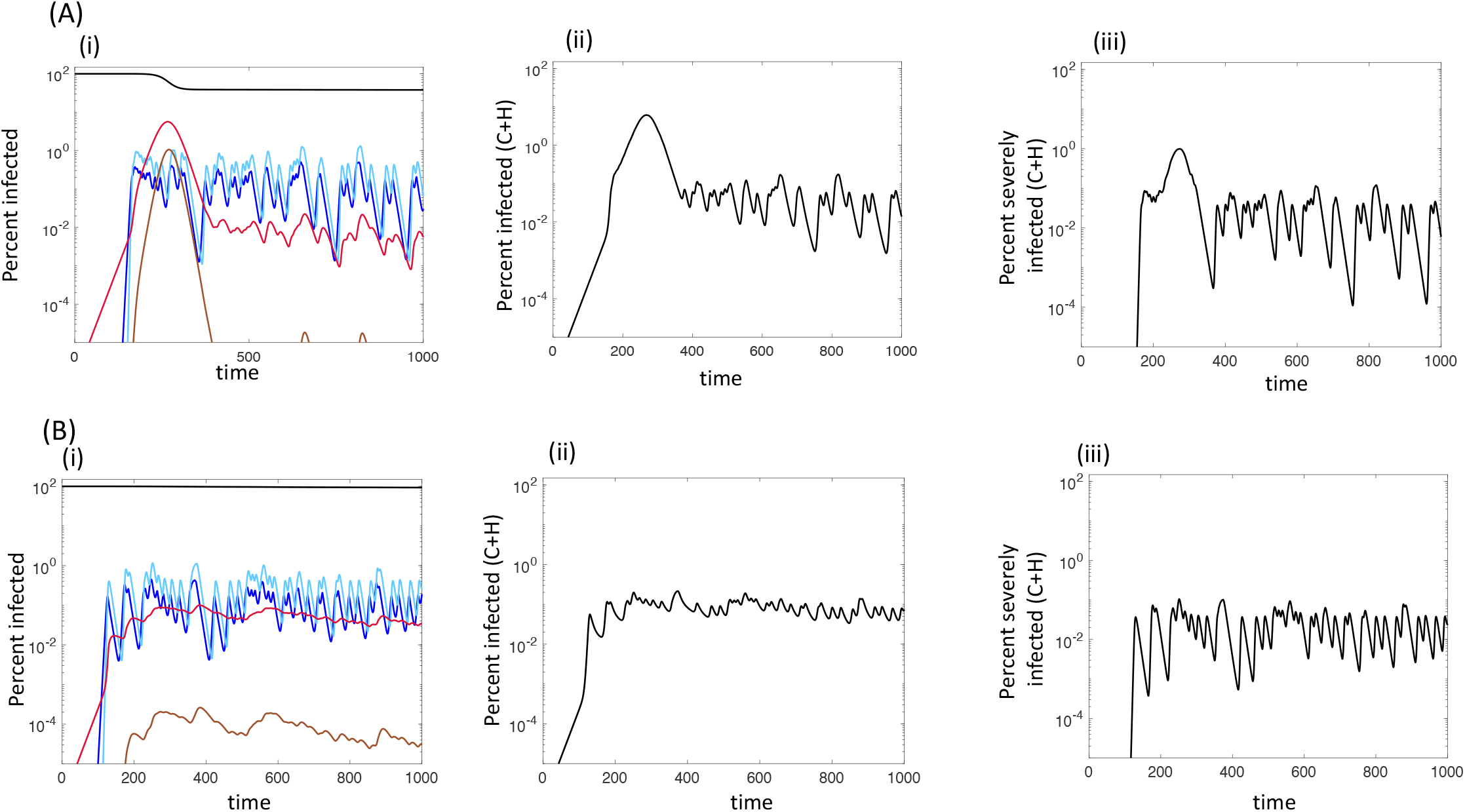
Model simulation assuming that in the community at large, R_0_=1.25, and in the hot zones, R_0_=3.75, and assuming that severely infected individuals have a higher rate of virus transmission than mildly infected individuals. (A) Natural infection spread without interventions. (Ai) Individual sub-populations. The percent of infected people are plotted. Black shows the percent of uninfected individuals in the community at large. Red and brown show mildly and severely infected individuals in the community at large. Dark and light blue show the percent of all mildly and severely infected individuals across all the hot zones. (Aii) Percent of all infections across the community at large and all hot zones, given by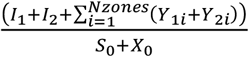. (Aiii) Percent of all severe infections across the community at large and all hot zones, given by 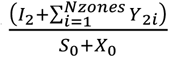. (B) Same kind of plots but with the implementation of non-pharmaceutical interventions in the community at large (rate of community infection β reduced 1.5 fold) when the percentage of infected individuals in the community at large reached 0.01%. (Bi) Percent individual sub-populations; (Bii) Percent of all infections across the community at large and all hot zones; (Biii) Percent of all severe infections across the community at large and all hot zones. Baseline parameters were given as follows. S_0_=1000, β_mild_=0.198×10^−3^, β_severe_=0.396×10^−3^, γ=1/7, f=0.1, X_0_=1, b_mild_=0.54, b_severe_=1.08, g=1/7, p=0.001, ξ=0.01, K_c_=400, K_h_=0.4, N_zones_=100.

Figure 4B shows that the dynamics during non-pharmaceutical interventions are qualitatively the same as in the previous version of the model. In this simulation, the intervention was initiated at relatively low infection levels in the community at large, while no interventions were put in place in hot zones. This again resulted in a prolonged infection plateau during the intervention, driven by continued hot zone transmission.

## 3. Modeling dynamic hot zones

So far we have considered hot zones as separate compartments/demes in which the same people repeatedly interact. These correspond most closely to nursing homes, hospitals, food processing plants, and prisons. There are, however, other hot zones settings in which people meet in conditions that facilitate virus spread, but where the same set of people do not meet repeatedly in the same location. Instead, a random set of people temporarily gathers in a hot zone, which subsequently dissolves and re-forms with a different set of people. Examples are restaurants, bars, or movie theaters. We refer to them as “dynamic hot zones” and here we explore a model that describes their effect on virus spread. We consider an agent-based model that tracks the fate of individual agents over time. The model assumes the existence of N individuals in a community in which the infection can spread. The individuals can be either uninfected, infected or recovered/dead. Virus spread in the general community is modeled as follows. The population is sampled repeatedly and randomly for updates. If the sampled individual is infected, virus transmission to a randomly chosen target individual is attempted with a probability P_inf_, and the individual recovers from the infection or dies with a probability P_remove_. If an infection is attempted, it is only successful if the randomly chosen target individual is uninfected. During a time step, the number of updates equals the number of infected individuals currently present in the population. For simplicity, we do not distinguish between mildly and severely infected individuals in this model.

In addition to virus spread in the general community, we assume that during each time step, a number of M hot zones form spontaneously, each consisting of N_hot_ individuals that are randomly drawn from the general population. Within each hot zone, infection transmission events occur according to the same algorithm as described above, but with an elevated transmission rate P^H^_inf_. During the next time step, all these hot zones dissolve and a new set of hot zones are generated with newly randomly drawn individuals from the community.

Simulation of this model results in a standard epidemiological infection wave without the observation of any of the atypical dynamics described in the previous sections. Figure 5A shows that the rate of infection spread is significantly faster in the presence of dynamic hot zone formation compared to its absence. Apart from that, however, the general shape of the dynamics is the same, and dynamics hot zone formation could be simply captured by standard SIR ordinary differential equations in which the average infection rate in the population is increased. Figures 5B and C show the dynamics assuming non-pharmaceutical interventions in the community (abolished community infectivity) and in the hot zones (abolished hot zone infectivity). A parameter regime is assumed in which either infection spread in the community alone or in hot zones alone can lead to a basic reproductive ratio of the virus that is greater than one. Therefore, when community interventions are implemented, the infection continues to grow for a while with a relatively slow rate until infection levels peak and decline (Figure 5B). Similar dynamics are observed if intervention occurs in the hot zones but not in the community (Figure 5C). In contrast to the compartment/deme models of hot zones in the previous sections, dynamic hot zones do not serve as a reservoir that can maintain prolonged infection persistence in the form of plateau dynamics. However, they do modulate the rate of progression of the outbreak.

**Figure 5.**
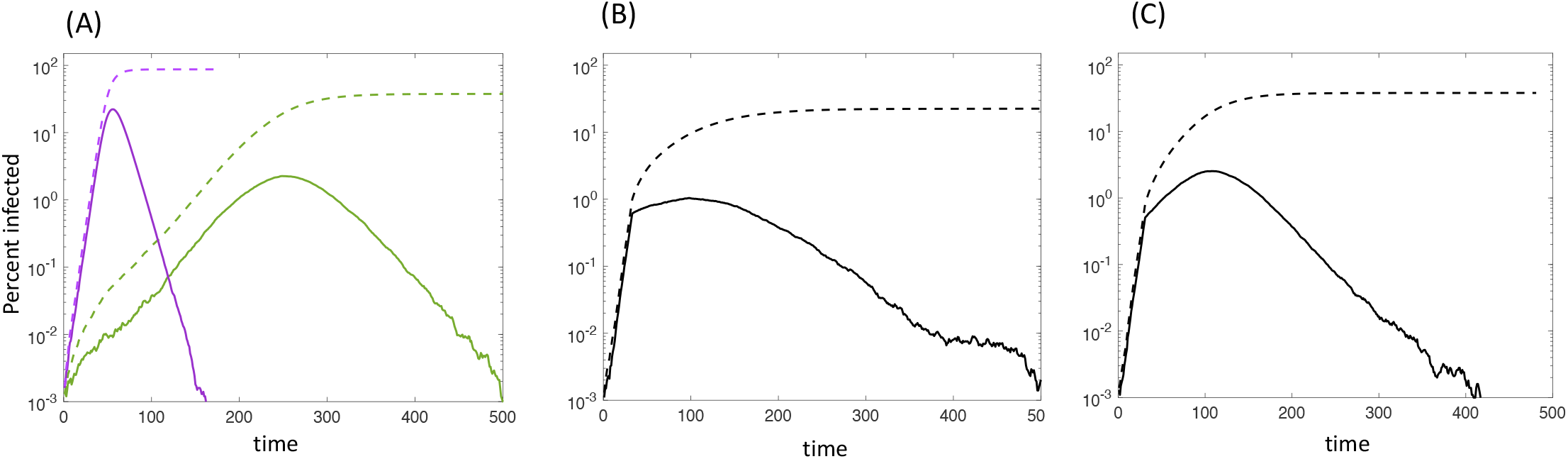
Computer simulations of the agent-based model with dynamics hot zones. Solid lines represent the percentage of infected individuals, and the dashed line show the cumulative percentage of infections. (A) Simulation in the absence of interventions. The purple line is a simulation that includes both community and hot zone transmission. The green line shows the dynamics in the absence of hot zone transmission. (B) Simulation of interventions in hot zones only. The virus transmission rate in the hot zone P^H^_inf_=0. (C) Simulation of interventions in the community only. The community virus transmission rate P_inf_=0. Baseline parameters were chosen as follows. P_inf_=0.1785 (with this value the community transmission rate yields an R_0_ about half of what has been reported for SARS-CoV-2); P^H^_inf_=0.714 (4 times larger than P_inf_); P_remove_=0.143; N=10^6^; M=10,000; N_hot_=25.

## 4. Discussion and Conclusion

Mathematical models have been used to characterize the dynamics of SARS CoV-2 and predict potential numbers of COVID-19 cases [21, 24-29], which has resulted in the estimation of the basic reproduction number [21, 22], a better understanding of expected transmission dynamics in the absence and presence of non-pharmaceutical interventions [30-37], and in the critical effect of age structure on disease dynamics [32, 38], among many other contributions [26]. Some of these models have been extremely useful for predicting and quantifying the demands on health care resources. The spread of SARS CoV-2, however, is characterized by a number of patterns that appear atypical when considered through the lens of such infection models [14, 39]. Individual atypical aspects have been accounted for by considering specific additions to infection models [14, 15, 39]. The static hot zone model presented here (where hot zones are described by individual compartments/demes/patches), however, can simultaneously give rise to a number of infection patterns that seem specific to SARS CoV-2. This could indicate that transmission in static hot zones is an important determinant of SARS CoV-2 dynamics, especially if it is assumed that the rate of virus spread in the community at large is relatively weak, and the rate of virus spread in hot zones is significantly faster.

(i) In accordance with data, the model predicts different growth phases of the pandemic as it progresses over time in individual geographical locations. The first phase is predicted to be characterized by slow growth, and consists almost entirely of mild cases. It is likely that in practice, this yields a silent phase of infection spread that is not detected by testing, because testing levels typically increase only once a sufficient number of symptomatic cases emerge. In agreement with this prediction, it is thought that in the US, SARS-CoV-2 first spread slowly without having been detected, before it was finally confirmed through testing. Also in agreement with model predictions, the earliest occurrence of severe cases (which are the ones most likely detected by testing) have been found in hot zones, in particular in nursing homes, e.g. in Washington State [40, 41]. Once virus spread in hot zones is initiated, the model predicts an acceleration of virus spread in the community at large, driven by the large reproductive potential of the virus in the hot zones. This is likely the point at which testing will start to document the disease spread, due to the emergence of severe cases in the community. Accordingly, epidemiological data showed a relatively fast increase of COVID19 cases when the pandemic was initially tracked in the US, characterized by a short doubling time of approximately 2-3 days. As the local epidemics progressed, however, data showed that the doubling times progressively slowed down, which is again in agreement with our modeling results. Once the infection has spread in the initial hot zones in the model, the hot zone dynamics enter a plateau phase, reflecting the balance between virus extinction in local hot zones and new hot zone seeding. This coincides with an increase in the predicted doubling time in the community at large because the plateau in the hot zone dynamics can no longer fuel continued fast community spread. These patterns also result in overall virus growth dynamics that are slower than exponential, a characteristic of relatively early COVID19 spread that has been seen in data across different countries [15].

(ii)It is an interesting modeling result that despite a low value of R_0_ assumed for the community at large, a fast temporary phase of community virus spread is predicted to occur that is driven by the relatively fast infection growth in the hot zones. This would most likely result in a significant over-estimation of the viral basic reproduction number in the community at large. This warrants further epidemiological investigations.

(iii) The model suggests that hot zones represent a reservoir for longer term virus persistence in the community at large, which is predicted to occur both in the absence and presence of non-pharmaceutical interventions. This manifests itself in a prolonged infection plateau after infection levels have peaked. Such infection plateaus have indeed been commonly found in epidemiological data on COVID19, and theoretical explanations have been offered that are based on behavioral dynamics [14] and network spread [39]. The model presented here adds to the list of possible explanations for the observed infection plateaus. Strikingly, our model suggests that a relatively late initiation of intervention measures results in a pronounced infection peak, and a subsequent sharp decline of infection levels before the plateau phase is reached, as has been observed for example in New York State. If interventions are put in place earlier, on the other hand, the model displays dynamics in which infection levels enter the plateau phase right away following the onset of interventions, without going through a clear peak, which has been found to be the case in several states, for example in California.

Regarding strategies to re-open society, the model makes interesting predictions about the types of non-pharmaceutical interventions that can be used to control the infection. In particular, the model points towards the importance of interventions that suppress virus spread in the hot zones. A basic result mentioned above is that hot zones represent a reservoir for long term virus persistence in the population. If hot zone transmission is suppressed during interventions alongside community transmission, this reservoir is eliminated, and the virus population would continuously decline towards extinction during the intervention phase as long as the reproduction number in the community is less than one. In this case, the dynamics would not converge to a long-lasting plateau. Further, if it is indeed true that R_0_ in the community is lower than that in the hot zones, it is possible that continued interventions in the hot zones, after interventions have been stopped in the community at large, could help reduce the extent of subsequent infection waves.

Our results add to previous work suggesting that the concept of R_0_ is complex in the context of SARS-CoV-2. Age and activity heterogeneity in mathematical models of of SARS-CoV-2 spread have been shown to lead to lower herd immunity thresholds [42]. A different degree of heterogeneity is present in our model, where a significantly different viral reproduction potential is postulated in hot zones and the community at large. The important finding in this respect is that despite an assumed low community R_0_, the documented rate of virus spread in the community can be temporarily fast, driven by the hot zones dynamics. If true, this has important implications for estimating the value of R_0_ in SARS-CoV-2 community infection.

Another important insight obtained from our models is that we can distinguish between two types of hot zones when considering infection dynamics, even though both types represent conditions/locations in which virus spread is facilitated. The first type are the “static” hot zones that are described by the deme or metapopulation model discussed above. The static hot zones can account for a variety of atypical infection dynamics patterns observed in COVID19 data, and they can provide a reservoir that can fuel longer term persistence of the infection in the form of infection plateaus. The central characteristic of such static hot zones is that a group of individuals remains there for prolonged or repeated periods and repeatedly interact (e.g. nursing homes, hospitals, prisons, etc). This is in contrast to “dynamic” hot zones, in which groups of people converge for a relatively short period of time (such as restaurants, bars, movie theaters), and where the same group of people does not meet repeatedly. Interestingly, such dynamic hot zones give rise to fundamentally different model properties compared to the static ones, and cannot account for any of the atypical infection dynamics patterns found in COVID19 data and cannot fuel the longer-term persistence of the virus at infection plateaus. However, these dynamic hot zones can modulate the rate of spread of the outbreak. Moreover, although mathematically undistinguishable from spread in the community at large, these dynamic hot zones are sensitive to different non-phamraceutical interventions than those required to target the transmission in the community at large, such as restricting access to dynamic hot zones versus large at home quarantines. This classification of infection hot zones into two groups (with different effects on infection dynamics) deserves further epidemiological investigation, especially in the context of determining what types of hot zones should be targeted by interventions to achieve maximal virus suppression.

## Data Availability

This work is based on mathematical models. No data are included.

## Notes

### Competing Interest Statement

The authors have declared no competing interest.

### Funding Statement

Support of grant NSF DMS 1662146/1662096 (NK & DW) is gratefully acknowledged. The authors or their institution never received payment or services from a third party for any aspect of the submitted work

