## Supplementary Materials for "Effect of hot zone infection outbreaks on the dynamics of SARS-CoV-2 spread in the community at large"

### Supplementary information

#### Contents

**1 Deterministic vs stochastic modeling** **1**

**2 A simplified ODE formulation for the model** **4**

2.1 Basic reproductive number calculations . . . . . . . . . . . . . 4

2.2 Solution structure . . . . . . . . . . . . . . . . . . . . . . . . . . 6

### 1 Deterministic vs stochastic modeling

Figure S1A presents simulations of the ODE model for static hot zones, given by the equations sets (1) and (2) in the main text. The total percent of infected individuals across the hot zones is shown in blue, and percent infection in the community at large in red. First consider the dynamics in the absence of non-pharmaceutical interventions (Figure S1(Ai)). Due to the deterministic nature of the model, all hot zones are immediately and simultaneously seeded, and significant virus growth across all hot zones is observed. This results in accelerated virus spread in the community at large. Following the peak, infection levels quickly decline to extinction due to herd immunity ion the model. Figure S1(Aii) simulates the same model when non-pharmaceutical interventions in the community at large are implemented (when percent infection levels in the community at large reach 0.1%). No interventions are assumed to occur in the hot zones. Upon start of the interventions, infection levels continue to grow in the community at large, due to the strong virus expansion that occurs in the hot zones. Infection percentages in the community at large, however, peak at lower levels compared to the simulation without interventions.

Figure S1(B) shows corresponding model predictions in a stochastic setting, by implementing Gillespie simulations of the ODE model, with identical parameters and initial conditions. Very different dynamics are observed, due to the stochastic seeding of the hot zones. First, consider the dynamics in the absence of interventions (Figure S1(Bi)) In contrast to the deterministic version of the model, not all hot zones are immediately and simultaneously seeded. Only some hot zones are initially seeded, but this still fuels accelerated virus spread in the community at large. Infection levels in individual hot zones go extinct in a relatively short amount of time (due to the small population size in each hot zone), but new hot zones are seeded. This balance between local infection extinction and seeding of new hot zones results in longer-term oscillatory dynamics in the percent infections in all hot zones, and this maintains oscillatory dynamics in the community at large as well. Therefore, in the stochastic version of the model, the hot zones serve as a reservoir that can maintain infection levels at a prolonged infection plateau. Figure S1(Bii) shows the dynamics when non-pharmaceutical interventions are assumed to occur in the community at large when the percent infected reaches 0.1%. Initiation of interventions results in a prolonged plateau phase of the dynamics, maintained by hot zone infections (where no interventions are assumed to occur).

We conclude that the stochastic seeding of hot zones significantly alters infection dynamics.

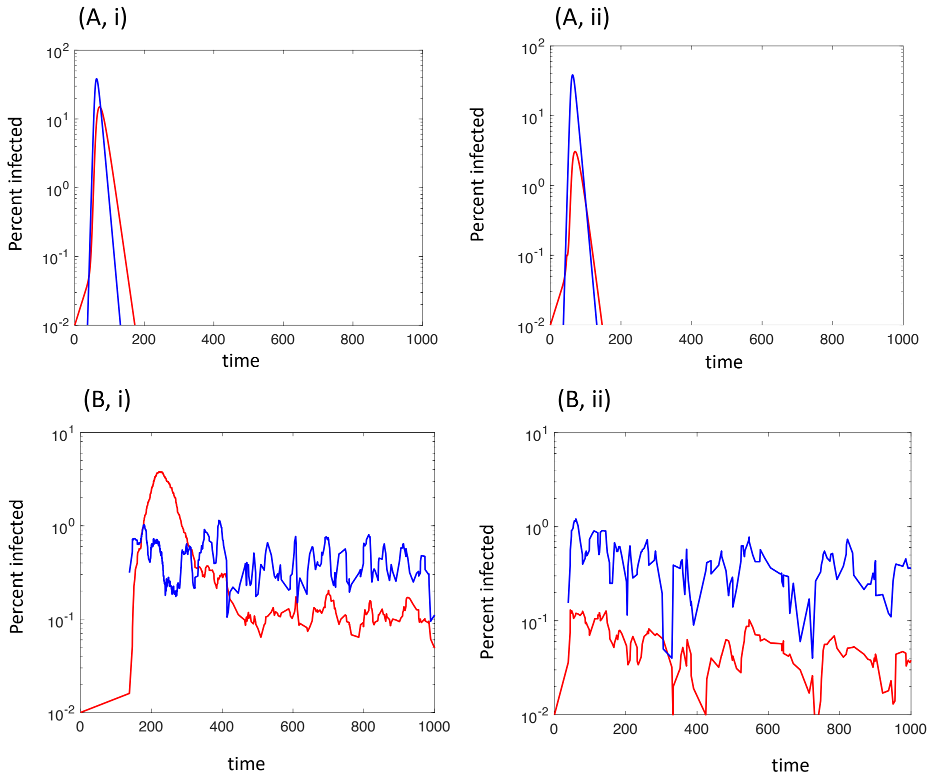


Figure S1: (A) Simulation of the ODE model, given by the equation sets (1) and (2) in the main text for the dynamics in the community at large (red) and the static hot zones (blue), respectively. (i) In the absence of non-pharmaceutical interventions. (ii) In the presence of non-pharmaceutical interventions, initiated when the infection percentage in the community at large reaches 0.1%. During interventions, the rate of infection in the community at large, $\beta$, is reduced 3-fold. No interventions occur in the hot zones. (B) Gillespie simulation of the same model with identical parameter values and initial conditions. (i) In the absence of non-pharmaceutical interventions. (ii) In the presence of non-pharmaceutical interventions. A single realization is shown for both cases. Parameters are given as follows. $S_0 = 100,000, \beta = 0.18 \times 10^{-5}, \gamma = 1/7, f = 1, X_0 = 100, b = 5.4 \times 10^{-3}, g = 1/7, h = 10^{-5}, \eta = 0.01, N_{zones} = 200$. For simplicity, this simulation did not distinguish between mild and severe infection, i.e. all infections are mild, $q_c = 0, q_h = 0$.

#### 2 A simplified ODE formulation for the model

Here we present mathematical analysis of a model similar to that described in the main text. Let us denote by $x_0, y_0$ the populations of susceptible and infected individuals in the community at large, and by $x_i, y_i$ with $1 \le i \le n$ the populations of susceptible and infected individuals in hot zone $i$. We have the following system:

$$
\begin{aligned}
\dot{x}_0 &= -(\beta_0 y_0 + \sum_{i=1}^{n} \tilde{\beta}_i y_i) x_0, & (1)\\
\dot{y}_0 &= (\beta_0 y_0 + \sum_{i=1}^{n} \tilde{\beta}_i y_i) x_0 - \gamma_0 y_0, & (2)\\
\dot{x}_i &= -(\beta_i y_i + \epsilon y_0/(y_0 + \xi)) x_i, \quad 1 \le i \le n & (3)\\
\dot{y}_i &= (\beta_i y_i + \epsilon y_0/(y_0 + \xi)) x_i - \gamma_i y_i, \quad 1 \le i \le n. & (4)
\end{aligned}
$$

Here, parameters $\gamma_i$ refer to recovery/remoal rates in different populations (subscript zero refers to the community at large $1 \le i \le n$ to the hot zones); $\beta_i$ is the infectivity parameter related to the infection process within a given population, $\tilde{\beta}_i$ is the infectivity between infecteds from hot zone $i$ and susceptibles in the community at large, and $\epsilon$ measures the (saturated) infection rate between the infected people in the community and susceptibles in the hot zones. There are two differences between the model analyzed here and the one in the main text:

- The model described here is simpler because it has a single population of infecteds for each of the locations and does not distinguishes between mildly and severely infected individuals.
- Within this reduced system, the parameters are somewhat less symmetric compared to the model in the main paper, such that we allow for parameter differences among the hot zones.

##### 2.1 Basic reproductive number calculations

To study the initial epidemic spread, we linearize the system around

$$x_0 = N_0, \quad y_0 = \bar{y}, \quad x_i = N_i, \quad y_i = 0, \quad 1 \le i \le n. \qquad (5)$$

Let us denote

$$\epsilon' = \frac{\epsilon}{xi}.$$

For the infection vector, $y = (y_0, y_1, \dots, y_n)^T$, we obtain the equation

$$\dot{y} = My,$$

where

$$M = \begin{pmatrix} \beta_0 N_0 - \gamma_0 & \tilde{\beta}_1 N & \dots & \tilde{\beta}_i N & \dots & \tilde{\beta}_n N \\ \epsilon' N_1 & \beta_1 N_1 - \gamma_1 & 0 & \dots & \dots & 0 \\ \epsilon' N_2 & 0 & \beta_2 N_2 - \gamma_2 & 0 & \dots & 0 \\ \dots & & & & & \\ \epsilon' N_i & 0 & \dots & \beta_i N_i - \gamma_i & \dots & 0 \\ \dots & & & & & \\ \epsilon' N_n & 0 & \dots & 0 & \dots & \beta_n N_n - \gamma_n \end{pmatrix}$$

If the characteristic polynomial of this matrix is given by $P(\lambda)$, then the equation $P(\lambda) = 0$ is equivalent to the following:

$$\beta_0 N_0 - \gamma_0 - \lambda = \sum_{j=1}^{n} \frac{\epsilon' \tilde{\beta}_j N_0 N_j}{\beta_j N_j - \gamma_j - \lambda} \equiv f(\lambda). \tag{6}$$

This equation can be used to solve for eigenvalues approximately in some regimes. Let us assume that

$$\epsilon' \ll |\beta_i N_i - \gamma_i - \beta_j N_j - \gamma_j|, \quad 0 \le i, j \le n, \quad i \neq j.$$

Then the right hand side of equation (6), $f(\lambda)$, is a piecewise continuous function of $\lambda$, with singularities at $\lambda = \beta_i N_i - \gamma_i$ with $1 \le i \le n$, and where this function satisfies

$$\lim_{\lambda \to (\beta_i N_i - \gamma_i)^+} f(\lambda) = -\infty, \qquad \lim_{\lambda \to (\beta_i N_i - \gamma_i)^-} f(\lambda) = +\infty.$$

This suggests that the roots of equation (6) will be near values $\beta_i N_i - \gamma_i$ with $0 \le i \le n$, and the distance from these values scales with $\epsilon'$. Therefore we can write down the eigenvalues as

$$\lambda_i = \beta_i N_i - \gamma_i + \delta_i, \quad \delta_i \sim \epsilon',$$

where $\delta_i$ is a small correction.

This calculation allows us to determine the parameter $R_0$ in this system. The epidemic does not grow from low numbers if all the eigenvalues are negative. Therefore, the condition for having a growing epidemic is that $\max_i \lambda_i > 0$. Let us suppose that

$$\max_i(\beta_i N_i - \gamma_i) = \beta_k N_k - \gamma_k.$$

Then, ignoring the correction, the condition $\lambda_k > 0$ is equivalent to $R_0 > 1$, where

$$R_0 = \frac{\beta_k N_k}{\gamma_k} + O(\epsilon').$$

This suggests that the $R_0$ value of the whole multi-population system is given by the maximum "individual" $R_0$ of one of its components.

#### 2.2 Solution structure

We can further find the solution structure and determine which eigenvalue defines the epidemic growth in the community ($y_0(t)$). Let us denote by $v^{(i)}$ the eigenvector associated with eigenvalue $\lambda_i$, $0 \leq i \leq n$. In the calculations below we will only include the lowest order terms in small $\epsilon'$. Eigenvector $v^{(0)}$ satisfies

$$(\lambda_i - \lambda_0)v_i^{(0)} = -\epsilon' N_i v_0^{(0)}, \quad 1 \leq i \leq n,$$

that is, $v^{(0)} = (1, \epsilon' N_1/(\lambda_0 - \lambda_1), \ldots, \epsilon' N_n/(\lambda_0 - \lambda_n))^T$. The first row of the matrix $M$ allows to calculate correction $\delta_0$:

$$\delta_0 = N\epsilon' \sum_{i=1}^{n} \frac{N_i}{\lambda_0 - \lambda_n}.$$

For components of $v^{(j)}$ with $j \neq 0$ we have

$$(\lambda_i - \lambda_0)v_i^{(j)} = -\epsilon' N_i v_0^{(j)}, \quad 1 \leq j \leq n, \quad i \neq j, \quad \delta_j v_j^{(j)} = \epsilon' N_j v_0^{(j)},$$

that is, setting $v_0^{(j)} = 1$, we have

$$v_i^{(j)} = \begin{cases} \frac{\epsilon' N_i}{\lambda_0 - \lambda_i}, & i \neq j, \\ \frac{\epsilon' N_j}{\delta_j}, & i = j. \end{cases}$$

and correction $\delta_j$ is determined from the first equation:

$$\delta_j = \frac{\epsilon' \tilde{\beta}_j N N_j}{\lambda_j - \lambda_0}.$$

Next we can determine the solution that corresponds to initial condition (5), by using the form

$$y(t) = \sum_{i=0}^{n} a_i v^{(i)} e^{\lambda_i t}.$$

We have $y(0) = (\bar{y}, 0, \ldots, 0)^T$. Writing out the components of this equation, we have

$$a_0 + \sum_{i=1}^{n} a_i = \bar{y}, \tag{7}$$

$$\frac{a_0 \epsilon' N_i}{\lambda_0 - \lambda_i} + \sum_{j \neq i} \frac{a_j \epsilon' N_i}{\lambda_0 - \lambda_i} + \frac{a_i \epsilon' N_i}{\delta_i} = 0, \quad 1 \leq i \leq n. \tag{8}$$

Let us use the following expansion for the coefficients: $a_i = A_i + B_i \epsilon'$, where $0 \leq i \leq n$ and $A_i, B_i$ are of order 1. Using only the lowest order terms in equation (8), we obtain $A_i = 0$ for all $1 \leq i \leq n$. Then from equation (7), we have $A_0 = \bar{y}$, and from equation (8),

$$B_i = \frac{\bar{y} \delta_i}{\lambda_i - \lambda_0},$$

which means that

$$a_0 = \bar{y} + O(\epsilon'), \quad a_i = \epsilon' \bar{y} \frac{\tilde{\beta}_i N N_i}{(\lambda_i - \lambda_0)^2}, \quad 1 \leq i \leq n.$$

In particular, the infection in the community is now given by

$$y_0(t) = \bar{y} \left( e^{\lambda_0 t} + \epsilon' N \sum_{i=1}^{n} \frac{\tilde{\beta}_i N_i}{(\lambda_i - \lambda_0)^2} e^{\lambda_i t} \right).$$

If the largest growth rate is $\lambda_k$ and $k \neq 0$, then the growth in the community will be influenced by the fast growth in the hot zone if

$$\epsilon' \gtrsim \frac{(\lambda_k - \lambda_0)^2}{\tilde{\beta}_k N_k N} e^{(\lambda_0 - \lambda_k) T},$$

where $T$ is the time scale relevant for the infection growth in the community. In other words, if the natural infection growth rate in the community is lower than that in some of the hot zones, the growth inside the hot zones can (temporarily) influence the growth in the community by increasing the speed of the infection spread there. The time-scale of this effect is related to the strength of the connection between the hot zone and the community.

This analysis can be generalized to the case where several values of $\lambda$ are the same (that is, several zones have the same $\beta_i N_i - \gamma_i$).
